## Appendix for "Using early detection data to estimate the date of emergence of an epidemic outbreak"

**3 Pseudo-algorithm** **12**

**References** **14**

### 1 Case datasets

#### 1.1 Alpha sequences reported in the UK

**Figure S1: Alpha sequences in the UK.** We apply our model to 406 sequenced samples (blue) collected up to November 11 (light-blue highlight) and reported by November 30, 2020 (date of submission to GISAID [1]). We do not consider samples collected between November 12 and November 30 to avoid the effects of reporting delays in the last days. For comparison, here we present the sequences reported after November 30, and up to December 15 (white), corresponding to a total of 455 sequenced samples.

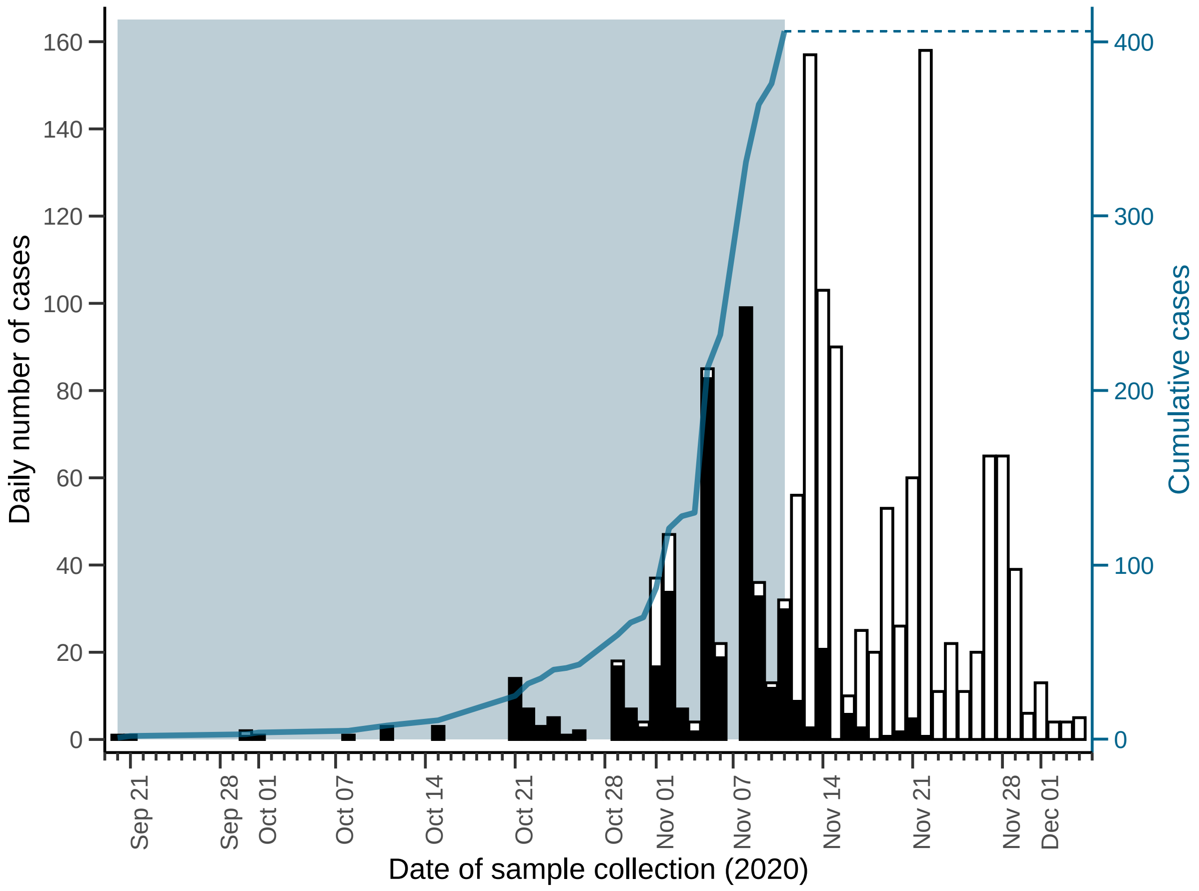

##### 1.1.1 GISAID data on Alpha cases in the UK

SUPPLEMENTAL TABLE

**Data Availability**

GISAID Identifier: EPI_SET_230104xg
doi: 10.55876/gis8.230104xg

All genome sequences and associated metadata in this dataset are published in GISAID's EpiCoV database. To view the contributors of each individual sequence with details such as accession number, Virus name, Collection date, Originating Lab and Submitting Lab and the list of Authors, visit 10.55876/gis8.230104xg

**Data Snapshot**

- EPI_SET_230104xg is composed of 409 individual genome sequences.
- The collection dates range from 2020-09-20 to 2020-11-11;
- Data were collected in 1 countries and territories;
- All sequences in this dataset are compared relative to hCoV-19/Wuhan/WIV04/2019 (WIV04), the official reference sequence employed by GISAID (EPI_ISL_402124). Learn more at https://gisaid.org/WIV04.

##### 1.2 Early cases of COVID-19 reported in Wuhan

**Figure S2: Early COVID-19 cases in Wuhan.** We apply our model to 3072 COVID-19 reported cases with symptoms onset by January 19, 2020 (blue). On January 20, the first public declaration of human-to-human transmission of the virus was made. Soon after that, the lockdown intervention was deployed nationally, along with mass testing, which explains the change in epidemic dynamics observed after that date (white).

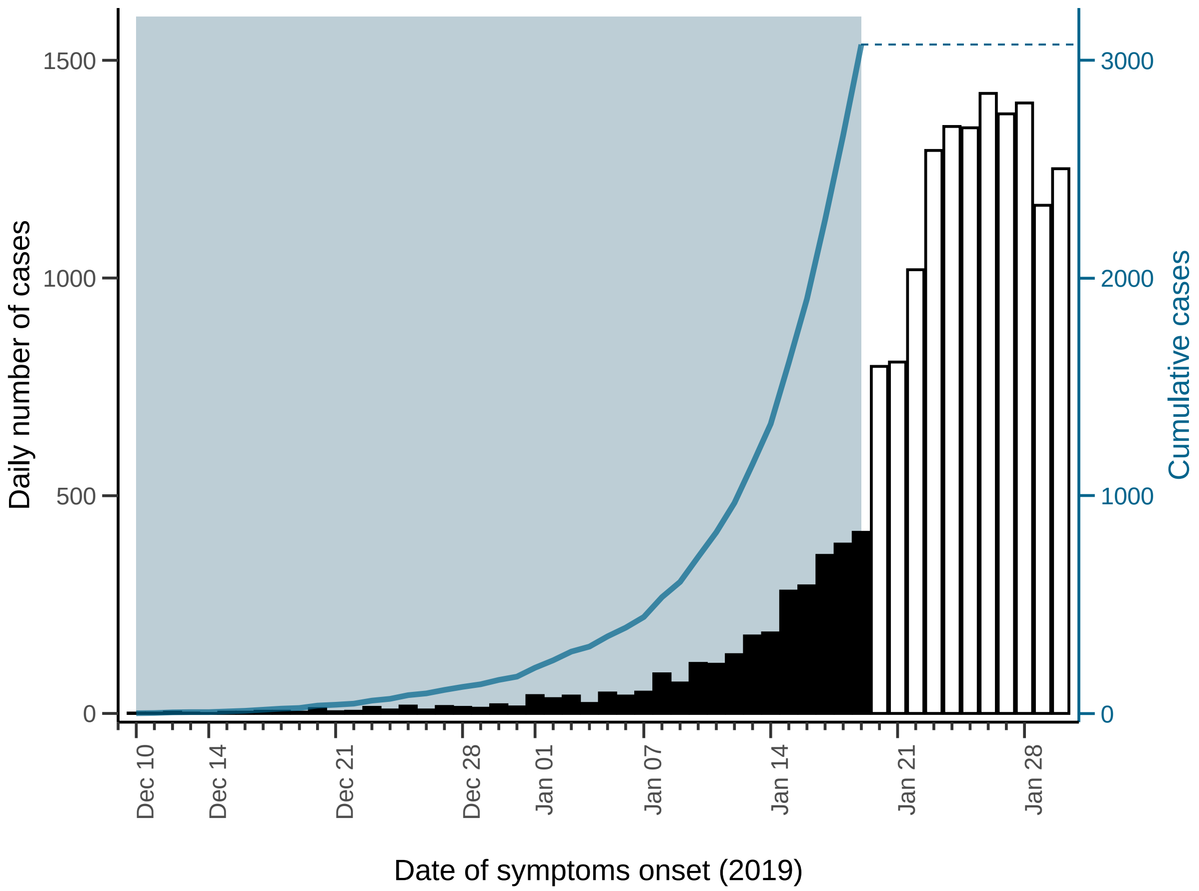

#### 2 Supplementary results

##### 2.1 Reanalysis of Roberts et al. (2021)

We used the same methodology as Roberts et al. (2021) [2] and ran it on updated datasets to assess the consequences of changes in the input case data. We found that the most recent datasets led to much later estimated dates of the first infection (see Figure S3).

**Figure S3: Reanalysis of Roberts et al. (2021) [2] with updated datasets.** The original analysis was done with Huang et al.'s dataset [3]. We re-ran the analysis on updated case datasets, using the same $N = 10$ number of case-days as in the original analysis.

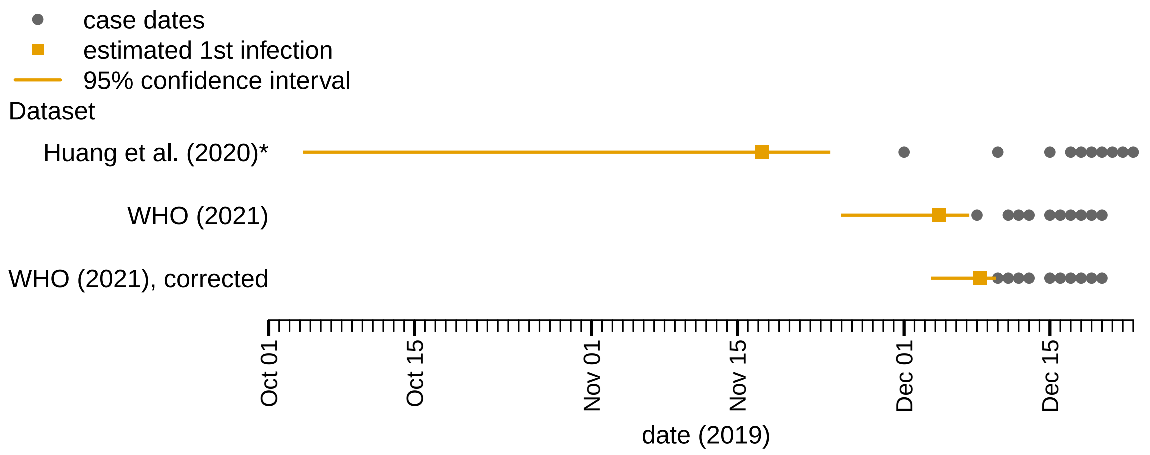

###### 2.1.1 Cumulative cases

**Figure S4: Cumulative cases of Alpha variant in the UK.** Accepted simulations (gray) and observed data (black).

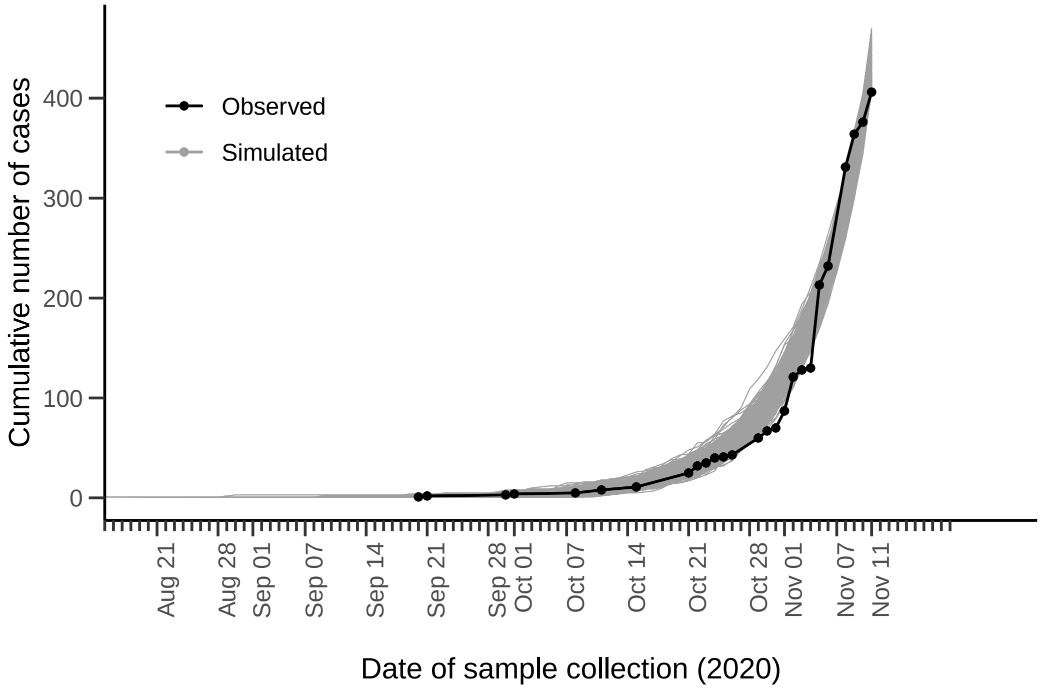

**Figure S5: Cumulative cases of COVID-19 in Wuhan.** Accepted simulations (gray) and observed data (black).

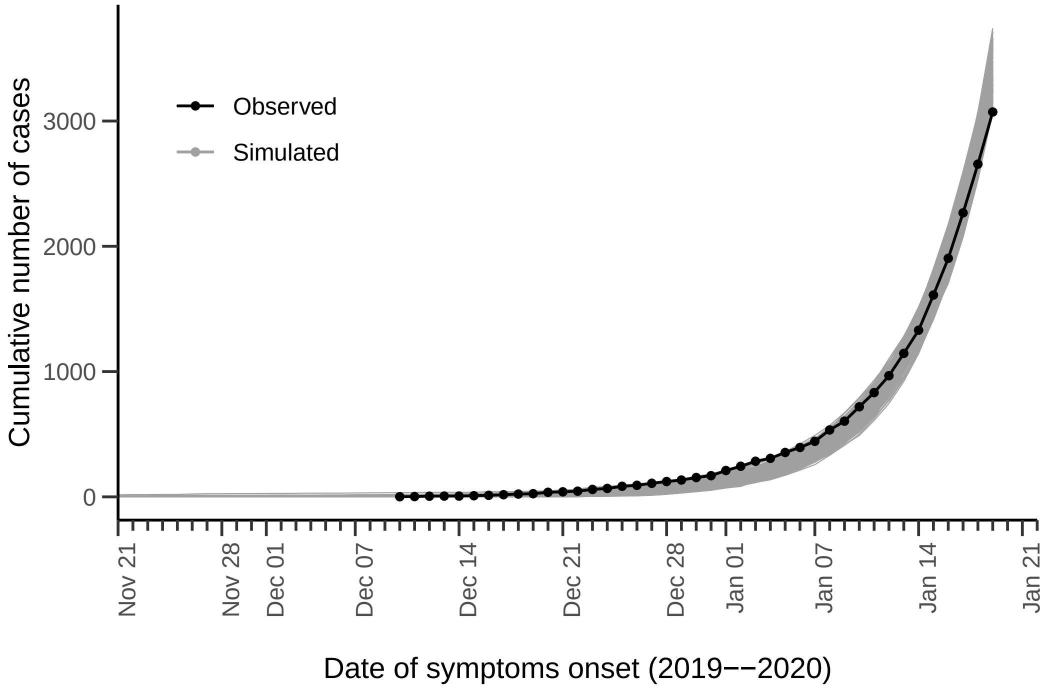

#### 2.2 Supplementary tables

##### 2.2.1 Model calibration

**Table S1: Model calibration.** Our results are obtained from a set of 5 000 simulations selected by the model calibration. Here, we summarize some metrics obtained by calibrating our model for the two applications, and show the observed data for comparison. The values and ranges of estimates correspond to medians and 95% interquantile ranges.

| | **Alpha** | | **COVID-19** | |
|---|---|---|---|---|
| **Metric** | **Observed** | **Estimated** | **Observed** | **Estimated** |
| Date of 1st case | Sep 20 | Sep 21 (Aug 24–Oct 4) | Dec 10 | Dec 8 (Nov 13–Dec 18) |
| Delay from $1^{st}$ to $N^{th}$ case (days) | 53 | 51 (38–79) | 41 | 42 (32–67) |
| Total cases at day of $N^{th}$ case | 406 | 432 (407–465) | 3072 | ~3 360 (3 080–3 690) |
| Proportion of accepted simulations | – | ~ 36% | – | ~ 14% |

##### 2.2.2 Estimates for the emergence date

**Table S2: Estimates for the emergence date.** Estimated dates of emergence. Median values and central 95% interquantile ranges (values between $2.5^{th}$ and the $97.5^{th}$ percentiles) are shown. Abbreviations: tMRCA= time of most recent ancestor.
* Simulations run using the model presented in [4], but with updated parameters matching ours.

| | **Method** | **Date of emergence** | **Earliest infection** | **Study** |
|---|---|---|---|---|
| **Alpha** | Population dynamics, based on $N$ observations. | Aug 21 (Jul 23–Sep 5), 2020 | Jun 13, 2020 | This study |
| | Population dynamics, based on the first observation. | Aug 19 (Jul 19–Sep 9), 2020 | Jun 2, 2020 | [4] updated* |
| | Phylodynamics estimating tMRCA. | Aug 28 (Aug 14–Sep 9), 2020 | Aug 6, 2020 | [5] |
| **COVID-19** | Population dynamics, based on $N$ observations. | Nov 28 (Nov 2–Dec 9), 2019 | Sep 26, 2019 | This study |
| | Phylodynamics model coupling transmission and viral genome evolution. | Nov 18 (Oct 16–Dec 6), 2019 | Sept 13, 2019 | [6] |

###### 2.2.3 Sensitivity analyses

**Table S3: Sensitivity analyses.** Estimated dates of emergence, obtained by varying the key epidemiological parameters (reproduction number, over-spreading and detection) as well as the parameters regarding the model calibration (i.e., the tolerances for simulation selection; see details in the Methods section of the main text). Median values and 95% interquantile ranges (values between 2.5th and the 97.5th percentiles) are shown. Baseline values are marked in boldface.

| | $R$ | $\kappa$ | $p_{\text{detect}}$ | $\theta_{\tau}$ | $\delta^{Y}_{\text{tol}}$ | **Date of emergence** |
|---|---|---|---|---|---|---|
| **Alpha** | 1.7 | 0.57 | 0.0105 | 12 | 0.3 | Aug 08 (Jul 06–Aug 26), 2020 |
| | **1.9** | **0.57** | **0.0105** | **12** | **0.3** | **Aug 21 (Jul 23–Sep 5), 2020** |
| | 2.1 | 0.57 | 0.0105 | 12 | 0.3 | Aug 29 (Aug 03–Sep 12), 2020 |
| | 1.9 | 0.35 | 0.0105 | 12 | 0.3 | Aug 23 (Jul 21–Sep 08), 2020 |
| | **1.9** | **0.57** | **0.0105** | **12** | **0.3** | **Aug 21 (Jul 23–Sep 5), 2020** |
| | 1.9 | 0.75 | 0.0105 | 12 | 0.3 | Aug 20 (Jul 18–Sep 03), 2020 |
| | 1.9 | 0.57 | 0.0050 | 12 | 0.3 | Aug 14 (Jul 18–Aug 29), 2020 |
| | **1.9** | **0.57** | **0.0105** | **12** | **0.3** | **Aug 21 (Jul 23–Sep 5), 2020** |
| | 1.9 | 0.57 | 0.0250 | 12 | 0.3 | Aug 28 (Jul 31–Sep 11), 2020 |
| | 1.9 | 0.57 | 0.0105 | 7 | 0.3 | Aug 23 (Jul 27–Sep 06), 2020 |
| | **1.9** | **0.57** | **0.0105** | **12** | **0.3** | **Aug 21 (Jul 23–Sep 5), 2020** |
| | 1.9 | 0.57 | 0.0105 | 15 | 0.3 | Aug 19 (Jul 23–Sep 02), 2020 |
| | **1.9** | **0.57** | **0.0105** | **12** | **0.3** | **Aug 21 (Jul 23–Sep 5), 2020** |
| | 1.9 | 0.57 | 0.0105 | 12 | 0.5 | Aug 20 (Jul 24–Sep 04), 2020 |
| **COVID-19** | 2.0 | 0.10 | 0.1500 | 6 | 0.3 | Nov 17 (Oct 20–Dec 02), 2019 |
| | **2.5** | **0.10** | **0.1500** | **6** | **0.3** | **Nov 28 (Nov 2–Dec 9), 2019** |
| | 3.5 | 0.10 | 0.1500 | 6 | 0.3 | Dec 05 (Nov 14–Dec 10), 2019 |
| | 2.5 | 0.05 | 0.1500 | 6 | 0.3 | Nov 30 (Nov 07–Dec 10), 2019 |
| | **2.5** | **0.10** | **0.1500** | **6** | **0.3** | **Nov 28 (Nov 2–Dec 9), 2019** |
| | 2.5 | 0.25 | 0.1500 | 6 | 0.3 | Nov 25 (Oct 29–Dec 05), 2019 |
| | 2.5 | 0.10 | 0.1000 | 6 | 0.3 | Nov 26 (Oct 31–Dec 08), 2019 |
| | **2.5** | **0.10** | **0.1500** | **6** | **0.3** | **Nov 28 (Nov 2–Dec 9), 2019** |
| | 2.5 | 0.10 | 0.2500 | 6 | 0.3 | Nov 30 (Nov 04–Dec 10), 2019 |
| | 2.5 | 0.10 | 0.1500 | 3 | 0.3 | Dec 01 (Nov 05–Dec 10), 2019 |
| | **2.5** | **0.10** | **0.1500** | **6** | **0.3** | **Nov 28 (Nov 2–Dec 9), 2019** |
| | 2.5 | 0.10 | 0.1500 | 12 | 0.3 | Nov 23 (Oct 28–Dec 05), 2019 |
| | 2.5 | 0.10 | 0.1500 | 6 | 0.1 | Nov 28 (Nov 04–Dec 10), 2019 |
| | **2.5** | **0.10** | **0.1500** | **6** | **0.3** | **Nov 28 (Nov 2–Dec 9), 2019** |
| | 2.5 | 0.10 | 0.1500 | 6 | 0.5 | Nov 28 (Nov 02–Dec 10), 2019 |

#### 2.3 Running our model on simulated data

Here, we ran our model on simulated case datasets and quantified the error of our estimates versus the *observed* date of emergence (observed in the simulation scenario and thus, known). We generated 100 simulated datasets and further ran our model, with 100 replicates for each combination, on subsets of different sizes; that is, we considered the case data up to the day of occurrence of the $N^{\text{th}}$ case, for increasing values of $N =\sim 1$, $N =\sim 10$ and $N =\sim 1\,000$.

**Figure S6: Distribution of the difference (in days) between the estimated date of the first infection and the actual date of the first infection in simulated data, for different values of $N$, the number of cases considered for the evaluation.** The vertical lines represent the median (full line) and the mean (dashed line) of the distributions.

a

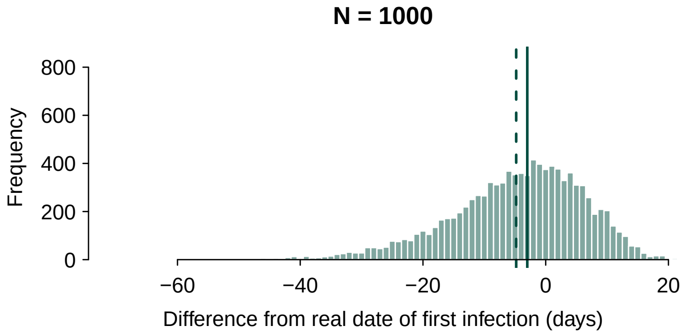

b

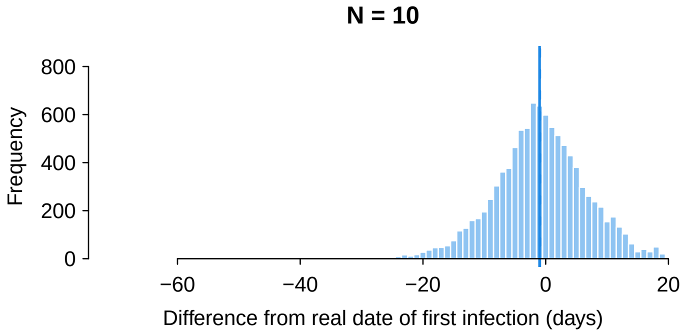

c

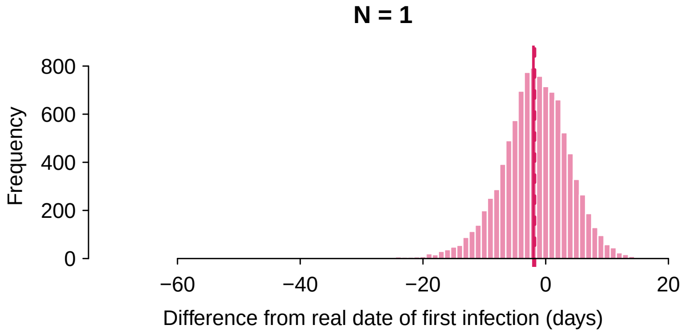

#### 2.4 Supplementary sensitivity analyses

##### 2.4.1 Using the first Alpha case only

**Figure S7: Estimates of the emergence date using information on the first case only.** Distributions of the emergence date for the Alpha variant in the UK, estimated using data ~~on data~~ on the first observed Alpha case (i.e., September 20, 2020) only, using our model (violet, top) and the model previously developed by (Czuppon et al., 2021) updating the parameters to match ours (red, middle; cf. the Methods section of the main text). We also plot the results from running our model on data of samples collected up to November 11, 2020 (i.e., our main results; blue, bottom), for comparison.

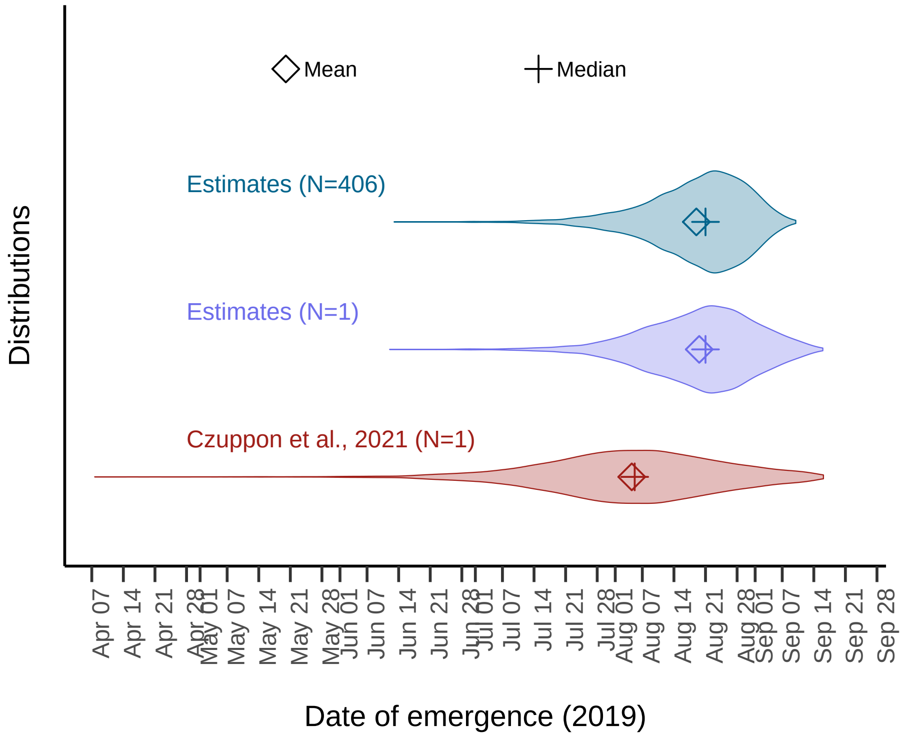

##### 2.4.2 Using other COVID-19 datasets

**Shorter dataset.** Here, we analyze the impact of applying our model to the dataset of COVID-19 cases (cf. Figure S2) with symptoms onset before December 31, 2019, the day of the first public declaration of the Wuhan cluster [7]. Based on the 169 reported cases of COVID-19 with symptoms onset before December 31, we estimate a date of emergence by November 24 (October 29–December 04), 2019, and not earlier than September 26, 2019.

**Anterior dataset.** In addition, we apply our model to case data published by the WHO in 2020 [8], which were later corrected [9]. The WHO data sets used in our model are adapted from the data sets available in a GitHub repository that summarizes published epidemic curves of early COVID-19 cases [10]. Results are summarized in Table S4.

**Figure S8: Estimates of the emergence date using truncated or outdated case datasets.** Distributions of the emergence date for the COVID-19 epidemic in Wuhan, estimated using data on cases with symptoms onset by December 31, 2019 (violet, middle) and by January 19, 2020 (i.e., our main results; blue, top), as well as an outdated, later corrected dataset (red, bottom). The first cases reported in the datasets are depicted by dashed lines colored accordingly.

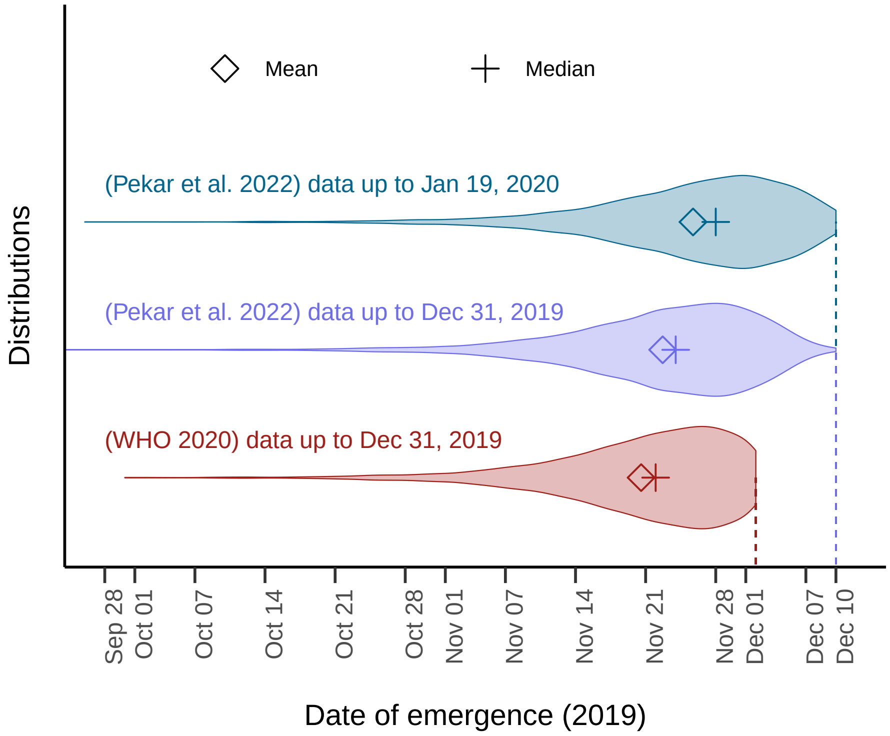

**Table S4: Impact of using different datasets.** Results obtained using different COVID-19 cases datasets. The estimated time elapsed between the first infection to the $N^{\text{th}}$ observed case yields the estimated date of outbreak emergence. In addition, we estimate the epidemic size at the date of detection of the $N^{\text{th}}$ case. The proportions of detected infections are retrieved for comparison with the input epidemic parameters (cf. Table 2 of the main text). Median and 95% interquantile ranges (i.e., values between 2.5$^{\text{th}}$ and the 97.5$^{\text{th}}$ percentiles) are shown, unless stated otherwise.

| | **Case dataset** | | |
|---|---|---|---|
| | **Pekar et. al 2022**[a] | **Pekar et. al 2022** | **WHO 2020** |
| **Observations** | | | |
| Date of $N^{\text{th}}$ case | Jan 19, 2020 | Dec 31, 2019 | Dec 31, 2019 |
| Date of first case | Dec 10 | Dec 10 | Dec 2 |
| Number of reported cases | 3 072 | 169 | 202 |
| **Simulations** | | | |
| Emergence date (2019) | Nov 28 (Nov 2–Dec 09) | Nov 24 (Oct 29–Dec 6) | Nov 22 (Oct 28–Dec 2) |
| Date of earliest infection | Sep 26, 2019 | Sep 24, 2019 | Sep 23, 2019 |
| Number of cases | ~3 360 (3 080–3 690) | 180 (169–210) | 220 (202–250) |
| Epidemic size[b] | ~63 400 (57 300–69 900) | ~3 500 (2 680–4 240) | ~4 160 (3 280–5 000) |
| Proportion of detected infections | 5.31% (5.06%–5.57%) | 5.3 (4.52–6.66) | 5.32 (4.55–6.49) |

[a] Dataset used in our baseline analyses.
[b] At day of infection of the $N^{\text{th}}$ case.

### 3 Pseudo-algorithm

The code was built in Julia and is available at a public Github repository: https://github.com/sjijon/estimate-emergence-from-data.

**Algorithm 1** Estimate the delay between the first infection and the $N^{\text{th}}$ case. Abbreviations: dist = Distribution.

**Require:** Time-series of the $N$ first observed cases, $\{d_i^{\text{obs}}\}_{i=1,\dots,K'}$
Context-specific epidemiological parameters (Distributions)
Lower bound for the number of infections ($I_{\text{min}}$)
Upper bound for the running time ($t_{\text{max}}$)
Number of simulations (num_sims)
Error tolerances ($\delta_{\text{tol}}$)

**Ensure:** Time elapsed between first infection and $N^{\text{th}}$ observed case
The simulated underlying epidemic (SimEpi)
The simulated cases (SimCases)

**procedure** READ DATA ▷ Returns ObsCases
    Read available data on cases
    Retrieve the number of observed cases, $N$, and their dates of occurence.
**end procedure**

**while** sim_num ≤ num_sims **do**

    **procedure** TRANSMISSION MODEL ▷ Returns SimEpi
        Initialize infections $I(t_0) = 1$
        **while** $t < t_{\text{max}}$ **do**
            Draw the number of secondary infections ▷ Negative binomial dist.
            **if** offspring was generated **then**
                Compute the time of those secondary infections, $\{t_i\}_{i=1,2,\dots}$ ▷ Gamma dist.
                Update number of infected at time of infection, $I(t)$
            **end if**
            Increase time step
        **end while**
        Compute daily number of infections, $I(d_k), k = 1, 2, \dots$
    **end procedure**

▷ ...Continues on the next page.

**Algorithm 1** (continuation)

```
    if I ≥ I_min then                                                      ▷ ~~Condition C1~~
        procedure DETECTION MODEL                                          ▷ Returns SimCases
            k=1
            for d_k, k = 1,2,... do
                Draw the daily number of detected infections among I(d_k)  ▷ Binomial dist.
                Compute detection time for each individual, {τ_i}_{i=1,...,M}  ▷ Gamma dist.
                Increase the daily number of cases, Y(d_j)
                if N^th case detected then
                    Keep only the detections occurring on the same day as N-th case
                    Exit loop
                end if
                k=k+1
            end for
        end procedure
    end if
    Identify the epidemic size at the day where the N^th case was infected

    if N_cases ≥ N then
        procedure COMPARE SIMCASES AND OBSCASES
            Condition C1: d_1 ≥ t_1
            ~~Condition C3:~~ (d_K^sim − d_1^sim)/(d_K'^obs − d_1^obs) ≤ δ_tol
            Condition C2: |Y^obs(d_k) − Y^sim(d_k)| ≤ δ'_tol N,    ∀k = K, K−1, K−2, ...
            if Conditions 2–4 hold then
                sim_num =+ 1                                               ▷ Select simulation
            end if
        end procedure
    end if

end while
```
